# Spatial Distribution and Factors Associated with Unmet Need for Family Planning Among Women of Reproductive Age in Zambia: Evidence from the 2024 Zambia Demographic and Health Survey

**DOI:** 10.64898/2026.11.02.26367140

**Authors:** Sibeso Philemon, Nasson Tembo, Choolwe Jacobs

## Abstract

Unmet need for family planning contributes to unintended pregnancies, unsafe abortions, and preventable maternal and child deaths, and remains high in sub-Saharan Africa. In Zambia, national estimates may conceal important geographic inequalities. We assessed the spatial distribution of unmet need for family planning among women of reproductive age in Zambia and the factors associated with it. We conducted a cross-sectional analysis of the 2024 Zambia Demographic and Health Survey (ZDHS), including 12,133 women aged 15-49 years. We used survey-weighted logistic regression, accounting for the stratified two-stage cluster design, to estimate adjusted odds ratios (AOR) with 95% confidence intervals (CI). Global spatial autocorrelation was assessed with Moran’s I, local hotspots with the Getis-Ord Gi* statistic, and spatial clusters with a purely spatial Bernoulli scan statistic in SaTScan. The weighted prevalence of unmet need was 27.0% (95% CI: 25.7-28.3%), ranging from 23.8% in Central to 31.6% in North-Western Province. Unmet need was spatially clustered (Moran’s I = 0.142, p = 0.018), with hotspots mainly in Lusaka, Southern, North-Western, and Northern provinces; SaTScan detected a primary cluster in Lusaka (observed-to-expected ratio 1.42, p < 0.001). Women in union (AOR 2.33, 95% CI: 1.90-2.86) and never-married women (AOR 2.20, 95% CI: 1.69-2.86) had higher odds of unmet need than formerly married women, as did women with five or more children compared with none (AOR 3.05, 95% CI: 2.25-4.13). Lower odds were observed among women aged 35-49 years (AOR 0.65, 95% CI: 0.52-0.83), women in the richest wealth quintile (AOR 0.62, 95% CI: 0.45-0.86), employed women (AOR 0.73, 95% CI: 0.64-0.84) and urban residents (AOR 0.80, 95% CI: 0.68-0.95). Unmet need for family planning in Zambia is high, geographically clustered and socially patterned. Family planning programmes should prioritise hotspot areas, particularly Lusaka, Southern and North-Western provinces, while targeting younger, higher-parity and poorer women.

## 1. Introduction

Family planning (FP) is a vital public health intervention that allows women and couples to decide freely whether, when, and how many children to have. By reducing unintended pregnancies and unsafe abortions, FP improves maternal and child health and contributes to gender equality and socio-economic development [1]. Unmet need for family planning refers to women who are fecund and sexually active, who wish to postpone or stop childbearing, but who are not using any method of contraception [2]. It is a key indicator for monitoring family planning programmes and for identifying barriers to contraceptive access and use.

In 2020, more than 218 million women in low- and middle-income countries were estimated to have an unmet need for contraception [3]. The burden is greatest in sub-Saharan Africa, where the prevalence of unmet need among married women ranges between 22.9% and 26.9% [4], driven by poverty, cultural norms, gender inequalities, and weak health systems [5]. In Zambia, despite steady progress in contraceptive use, about 20% of married women had an unmet need for family planning in the 2018 Zambia Demographic and Health Survey (ZDHS) [2]. This persisting gap hinders progress towards Sustainable Development Goal (SDG) target 3.7, which calls for universal access to sexual and reproductive health services by 2030.

Unmet need is not evenly distributed within countries. In Zambia, women in rural and hard-to-reach areas face longer distances to health facilities, fewer trained staff, and less exposure to family planning information [6], and provinces such as Luapula and Eastern have previously reported higher levels of unmet need [7]. Spatial analyses in Uganda and Ethiopia have shown that unmet need concentrates in identifiable geographic hotspots, allowing resources to be directed to high-burden areas [8, 9]. Previous Zambian studies, however, have relied on national and provincial estimates from non-spatial regression models [2, 3, 7], and national averages can mask local inequalities. To our knowledge, formal spatial statistics have not yet been applied to geo-referenced data from the 2024 ZDHS to locate clusters of unmet need in Zambia.

This study therefore aimed to assess the spatial distribution of unmet need for family planning and to determine the factors associated with unmet need among women of reproductive age (15-49 years) in Zambia, using data from the 2024 ZDHS.

## 2. Materials and methods

### 2.1 Study area and setting

The study analysed data from all ten provinces of Zambia: Central, Copperbelt, Eastern, Luapula, Lusaka, Muchinga, Northern, North-Western, Southern and Western. Zambia is a landlocked country in southern Africa with a population of approximately 20 million people. The provinces differ substantially in population density, urbanisation, economic development and access to health services.

### 2.2 Study design, data source and study population

This was a cross-sectional study, constituting a secondary analysis of the 2024 ZDHS. The ZDHS is part of the global Demographic and Health Surveys (DHS) Program and provides nationally representative population and health indicators [10]. The 2024 ZDHS used a stratified two-stage cluster sampling design. In the first stage, 545 enumeration areas (EAs) were selected from the national master sampling frame with probability proportional to EA size, after stratification by province and by urban or rural residence (20 sampling strata). In the second stage, a fixed number of households were selected by systematic random sampling from an updated household listing in each selected EA. All women aged 15-49 years who were usual residents of, or had slept the previous night in, the selected households were eligible for interview with the Women’s Questionnaire. Data were collected between January and July 2024, and access to the data set was granted and accessed on 15th December ,2025.Geographic coordinates of EA centroids, released by the DHS Program with random displacement for confidentiality, were used for the spatial analyses.

For this study, we extracted the pertinent variables from the women’s individual recode (IR) file of the 2024 ZDHS. A total of 13,951 women aged 15-49 years were interviewed in the 2024 ZDHS, and 12,133 women formed the analytic sample

### 2.3 Study variables

#### Outcome variable

The outcome was unmet need for family planning, derived from the DHS variable v626a and coded as a binary variable (1 = unmet need for spacing or limiting births, i.e., v626a codes 1 and 2; 0 = all other codes, including women using contraception). Consistent with the standard DHS definition, unmet need was present among fecund, sexually active women who wished to delay or limit childbearing but were not using any contraceptive method.

#### Explanatory variables

Explanatory variables were selected using an integrated conceptual framework drawing on Andersen’s Behavioral Model of Health Services Use [11] and the Social Ecological Model [20]. Predisposing factors were age group (15-24, 25-34, 35-49 years), highest educational level (no education, primary, secondary, higher), and marital status (never married, in union, formerly married). Enabling factors were household wealth quintile (poorest to richest), employment status (working, not working), place of residence (urban, rural), province, and whether distance to a health facility was reported as a problem (yes, no). Need factors were parity, measured as the number of living children (0, 1-2, 3-4, 5 or more), and fertility preference (DHS variable v602, entered as a numeric code). Exposure to family planning messages in the media and ideal number of children were also considered but were not retained in the final model

### 2.4 Data management

Data were imported into R from Stata format using the haven package, and variable names were standardised with the janitor package. Continuous age was grouped into categories, marital status was recoded into three groups, and the number of living children into four groups. Sampling weights (v005) were divided by 1,000,000 in accordance with DHS guidance. Two candidate variables had substantial missing data, partner’s fertility desire (75.3%) and women’s decision-making autonomy (46.5%), and were excluded from all models. Women with missing values on any other model variable were excluded (complete-case analysis). A sensitivity analysis that retained these two variables with missing-category indicators was performed.

### 2.5 Statistical analysis

#### 2.5.1 Descriptive analysis

Weighted frequencies and proportions were computed for all study variables. The weighted prevalence of unmet need and its 95% confidence interval (CI) were estimated nationally and by province. Survey-weighted chi-square tests were used to assess bivariable associations between each explanatory variable and unmet need.

#### 2.5.2 Factors associated with unmet need

A complex survey design object specifying clusters, strata, and sampling weights was created with the R survey package (svydesign), and survey-weighted generalised linear models (svyglm) with a quasibinomial family were fitted. Univariable models were estimated first, and variables with p < 0.20 were entered into the multivariable model. Crude odds ratios (COR) and adjusted odds ratios (AOR) with 95% CI are reported, and statistical significance was set at p < 0.05. Multicollinearity was assessed with variance inflation factors (VIF), with VIF > 10 indicating problematic collinearity [12]. Because women were sampled in clusters, all estimates accounted for the survey design: clustering by EA and stratification were specified in the design object, so that standard errors and confidence intervals reflect the two-stage cluster sampling rather than assuming independent observations.

#### 2.5.3 Spatial analysis

The EA-level prevalence of unmet need was linked to the geographic coordinates of EA centroids. Global spatial autocorrelation was tested with Moran’s I, using a k-nearest-neighbour spatial weights matrix (k = 5). Local clusters of high values (hotspots) and low values (coldspots) were identified with the Getis-Ord Gi* statistic at the 95% (|z| ≥ 1.96) and 99% (|z| ≥ 2.58) confidence levels. Maps were produced with the sf and tmap packages in R version 4.4.1.

As a confirmatory approach based on a different algorithm, spatial scan statistics were applied in SaTScan version 10.1.3 using a purely spatial Bernoulli model, with women with unmet need as cases and women without unmet need as controls. A circular scanning window with a maximum size of 50% of the population at risk was used, and p-values for the log-likelihood ratio (LLR) were derived from 999 Monte Carlo replications. Clusters with p < 0.05 were considered statistically significant and were mapped in QGIS version 3.42.

### 2.6 Ethical considerations

The 2024 ZDHS protocol was approved by the University of Zambia Biomedical Research Ethics Committee (UNZABREC) and the ICF Institutional Review Board, and all respondents provided informed consent before the interview. For this secondary analysis, ethical clearance and a waiver of individual informed consent were obtained from UNZABREC (Ref. No. 7631-2026; 16 February 2026), and authority to conduct the study was granted by the National Health Research Authority (NHRA; Ref. No. NHRA-3280/19/02/2026; 2 March 2026). Access to the de-identified dataset was granted by the DHS Program under its standard data use agreement. No information that could identify individual respondents or households was used.

## 3. Results

### 3.1 Characteristics of the study population

A total of 12,133 women aged 15-49 years were included in the analysis (Table 1). Most women lived in rural areas, were currently in a marital union and had at least one living child, and close to half had attained secondary education. More than half of the women (55.9%) were employed, and 39.8% reported that distance to a health facility was a problem.

**Table 1.** Weighted sociodemographic characteristics of women aged 15-49 years, Zambia, 2024 ZDHS (n = 12,133)

| **Characteristic** | **Category** | **n (%)** |
|---|---|---|
| Age group | 15-24 years | 5,154 (42.5%) |
|  | 25-34 years | 3,604 (29.7%) |
|  | 35-49 years | 3,375 (27.8%) |
| Education | No education | 787 (6.5%) |
|  | Primary | 5,050 (41.6%) |
|  | Secondary | 5,435 (44.8%) |
|  | Higher | 861 (7.1%) |
| Residence | Urban | 5,405 (44.5%) |
|  | Rural | 6,728 (55.5%) |
| Wealth index | Poorest | 2,378 (19.6%) |
|  | Poorer | 2,334 (19.2%) |
|  | Middle | 2,498 (20.6%) |
|  | Richer | 2,432 (20.0%) |
|  | Richest | 2,491 (20.5%) |
| Parity (living children) | 0 | 3,486 (28.7%) |
|  | 1-2 | 3,778 (31.1%) |
|  | 3-4 | 2,670 (22.0%) |
|  | 5 or more | 2,199 (18.1%) |
| Marital status | Never married | 4,132 (34.1%) |
|  | In union | 6,285 (51.8%) |
|  | Formerly married | 1,716 (14.1%) |
| Employment status | Not working | 5,351 (44.1%) |
|  | Working | 6,782 (55.9%) |
| Distance to facility a problem | No | 7,304 (60.2%) |
|  | Yes | 4,829 (39.8%) |
| Unmet need for FP | No unmet need | 8,857 (73.0%) |
|  | Unmet need present | 3,276 (27.0%) |

n (%) = weighted count (weighted percentage), derived from survey-weighted analysis using the complex survey design (svydesign). FP = family planning.

### 3.2 Prevalence and spatial distribution of unmet need

The weighted national prevalence of unmet need for family planning was 27.0% (95% CI: 25.7-28.3%). Prevalence was highest in North-Western (31.6%), Lusaka (31.4%), and Southern (30.9%) provinces and lowest in Central Province (23.8%) (Table 2; Fig 1).

**Table 2.** Distribution of women aged 15-49 years by province and unmet need for family planning, Zambia, 2024 ZDHS.

| **Province** | **No unmet need, n (%)** | **Unmet need, n (%)** | **p-value** |
|---|---|---|---|
| Central | 1,454 (12%) | 171 (9.4%) | <0.001 |
| Copperbelt | 1,408 (12%) | 157 (8.6%) |  |
| Eastern | 1,368 (11%) | 202 (11%) |  |
| Luapula | 1,126 (9.3%) | 135 (7.4%) |  |
| Lusaka | 1,496 (12%) | 225 (12%) |  |
| Muchinga | 874 (7.2%) | 139 (7.6%) |  |
| Northern | 1,119 (9.2%) | 188 (10%) |  |
| North-Western | 981 (8.1%) | 186 (10%) |  |
| Southern | 1,345 (11%) | 257 (14%) |  |
| Western | 962 (7.9%) | 158 (8.7%) |  |

**Fig 1.**
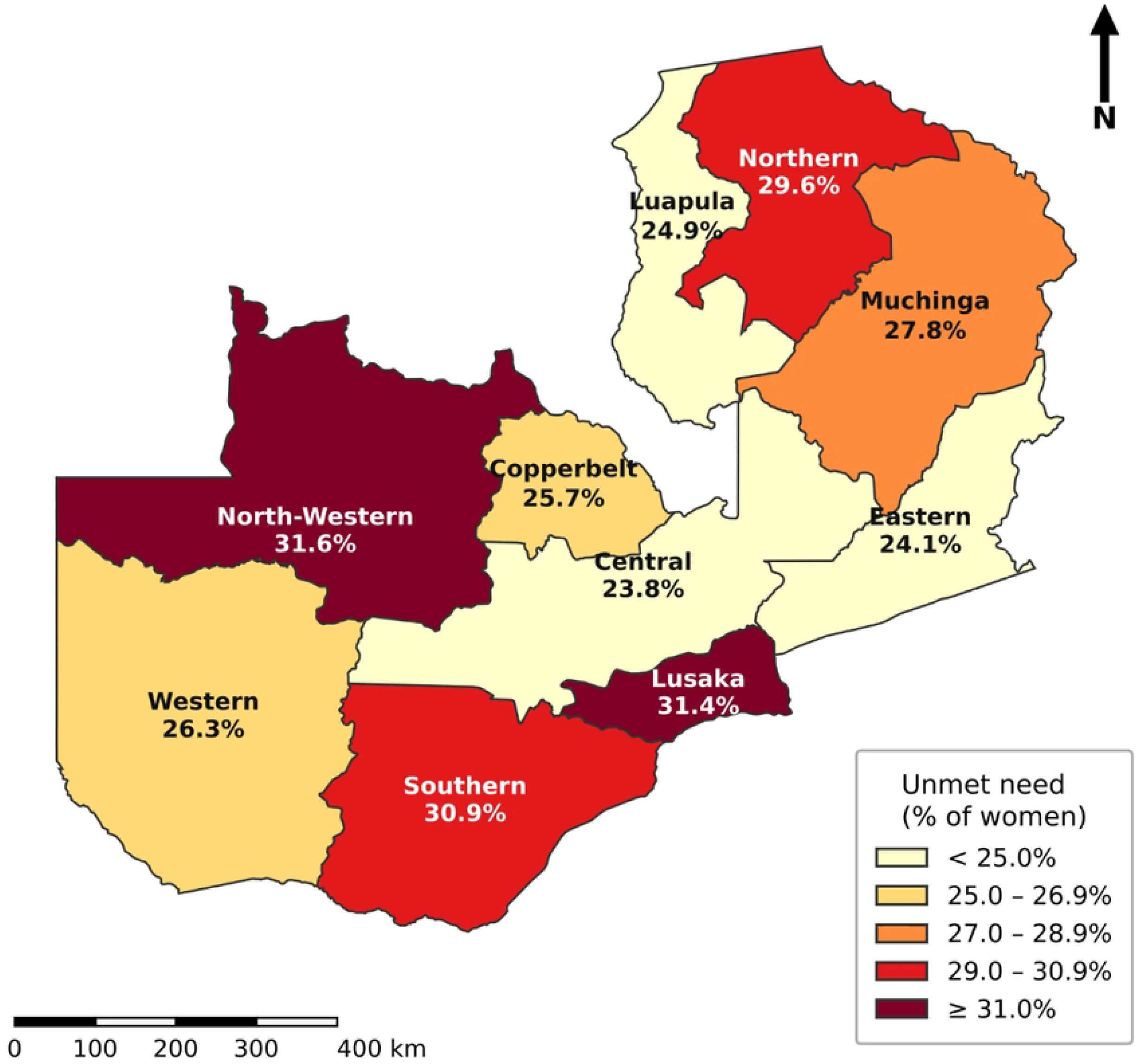
Provincial distribution of unmet need for family planning among women aged 15-49 years, Zambia, 2024 ZDHS. Provincial values are the weighted prevalence estimates in Table 2. Province boundaries: Natural Earth (public domain).

Unmet need showed statistically significant positive global spatial autocorrelation across the 545 EAs (Moran’s I = 0.142; z-score = 2.43; p = 0.018).

In the Getis-Ord Gi* analysis, 27 of the 545 EAs (5.0%) were statistically significant hotspots: 14 (2.6%) at the 99% confidence level, located in Lusaka, Southern and North-Western provinces, and 13 (2.4%) at the 95% level, located in Northern, Lusaka and Southern provinces. Fifteen EAs were coldspots: two (0.4%) at the 99% level in Eastern and Western provinces and 13 (2.4%) at the 95% level in Central and Eastern provinces. The remaining 503 EAs (92.3%) were not significant (Fig. 2).

**Fig 2.**
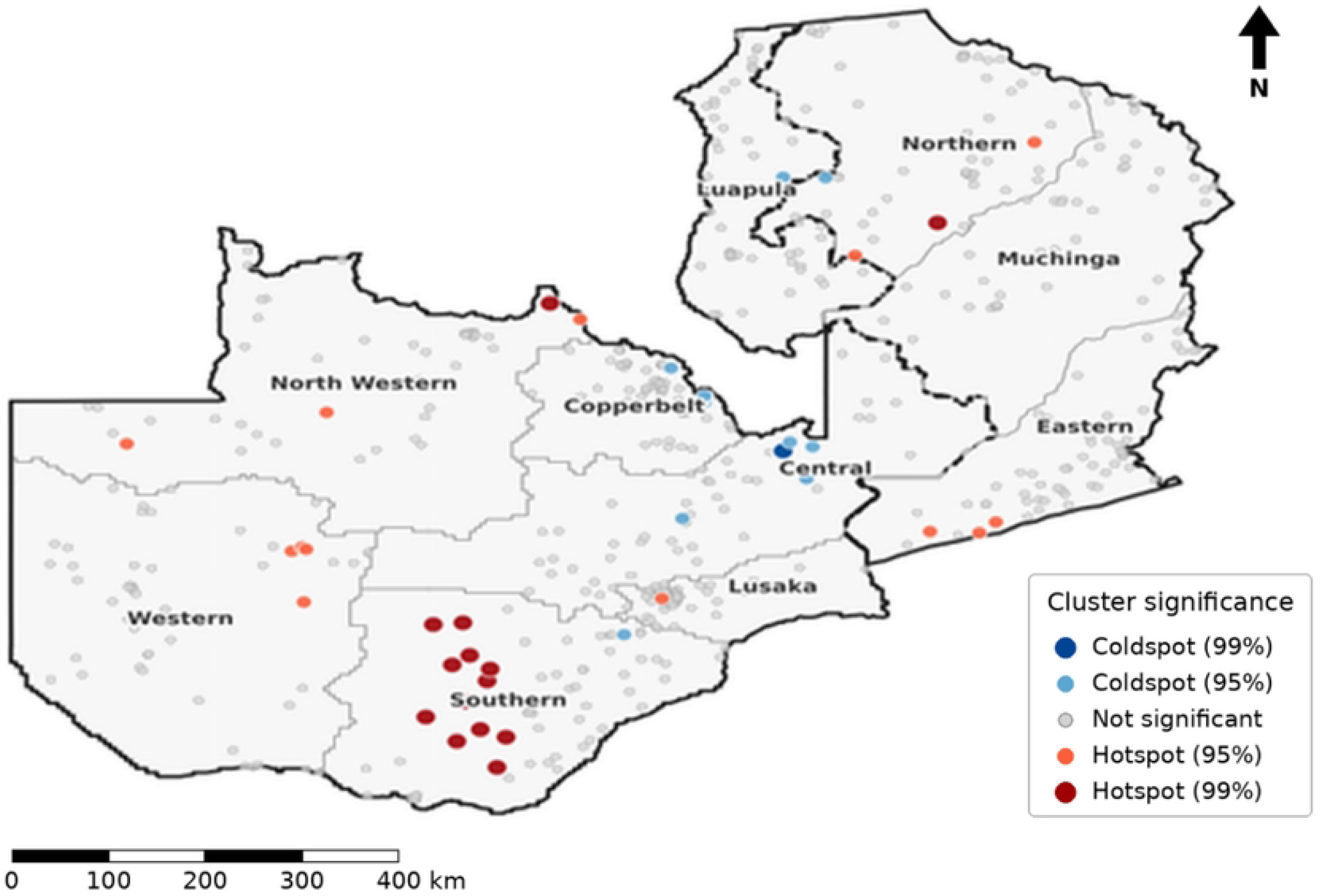
Hotspot analysis (Getis-Ord Gi*) of unmet need for family planning by enumeration area among women aged 15-49 years, Zambia, 2024 ZDHS

The SaTScan analysis identified three statistically significant clusters of elevated unmet need (Table 3; Fig 3). The primary cluster comprised 27 EAs in Lusaka Province (observed-to-expected ratio 1.42; LLR = 38.42; p < 0.001). Secondary clusters were located in Southern Province (18 EAs; observed-to-expected ratio 1.31; p = 0.003) and North-Western Province (14 EAs; observed-to-expected ratio 1.28; p = 0.021).

**Table 3.** Spatial clusters of unmet need for family planning detected by the purely spatial Bernoulli scan statistic, Zambia, 2024 ZDHS.

| **Cluster** | **Location (province)** | **No. of DHS clusters** | **Observed cases (n)** | **Expected cases** | **Obs/Exp ratio** | **LLR** | **p-value** |
|---|---|---|---|---|---|---|---|
| Primary | Lusaka | 27 | 1,842 | 1,297.9 | 1.42 | 38.42 | <0.001 |
| Secondary 1 | Southern | 18 | 1,103 | 841.5 | 1.31 | 18.76 | 0.003 |
| Secondary 2 | North-Western | 14 | 827 | 645.4 | 1.28 | 12.94 | 0.021 |

SaTScan™ version 10.1.3; purely spatial Bernoulli model; maximum spatial cluster size = 50% of the population at risk; Monte Carlo replications = 999. LLR = log-likelihood ratio. Clusters significant at p < 0.05 are reported. Geographic coordinates are DHS cluster centroids, displaced by the DHS Program by up to 2 km in urban and up to 5 km in rural areas (up to 10 km for 1% of rural clusters).

**Fig 3.**
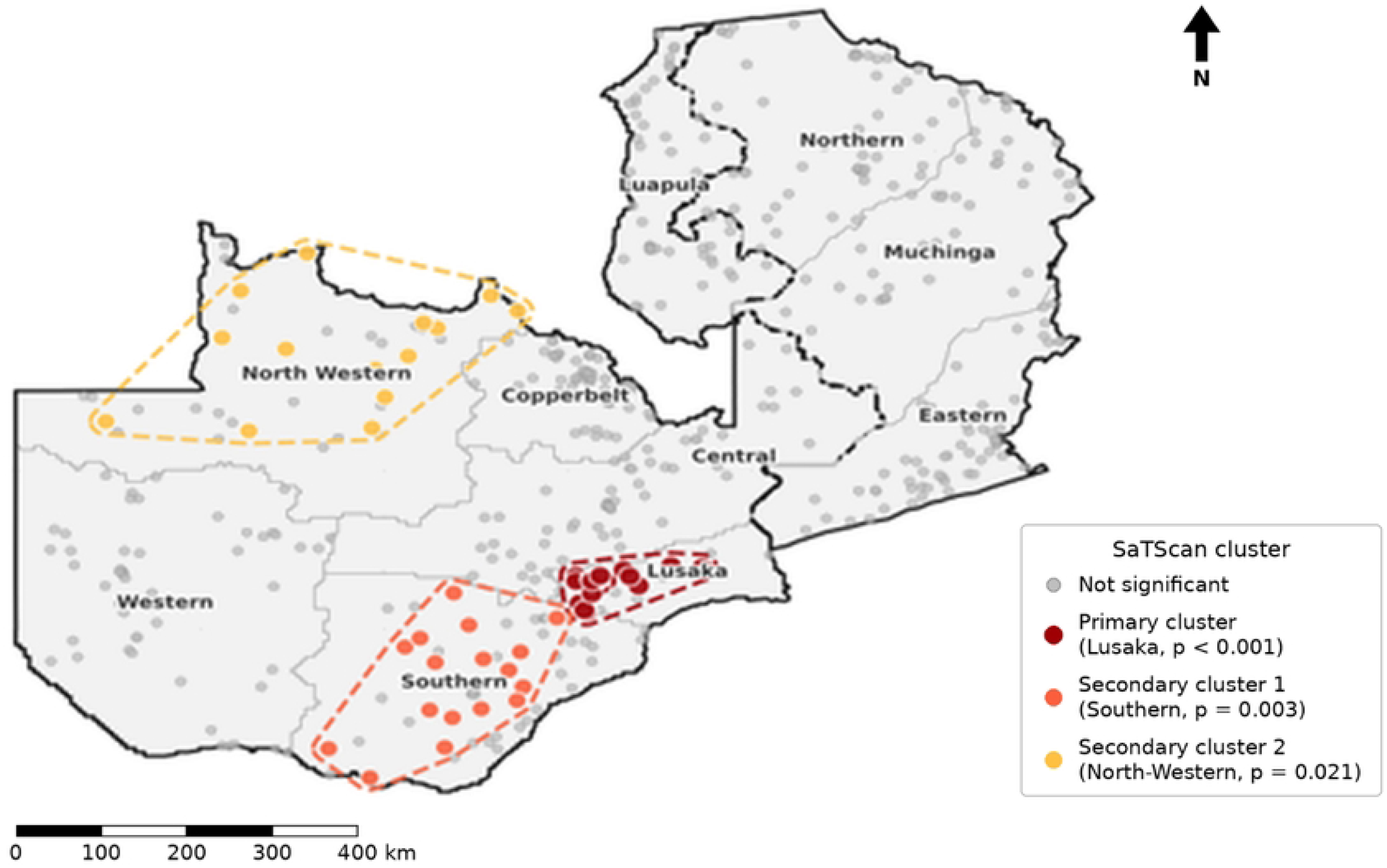
Spatial clusters of unmet need for family planning identified by SaTScan among women aged 15-49 years, Zambia, 2024 ZDHS

### 3.3 Factors associated with unmet need for family planning

Table 4 presents the crude and adjusted odds ratios. No multicollinearity was detected (all VIF < 3.0). In the adjusted model, marital status and parity were the strongest correlates of unmet need. Women currently in union (AOR 2.33, 95% CI: 1.90-2.86) and never-married women (AOR 2.20, 95% CI: 1.69-2.86) had higher odds of unmet need than formerly married women. The odds increased with the number of living children, from AOR 2.04 (95% CI: 1.64-2.54) for 1-2 children to AOR 3.05 (95% CI: 2.25-4.13) for five or more children, compared with women with no children.

**Table 4.** Survey-weighted logistic regression of factors associated with unmet need for family planning among women aged 15-49 years, Zambia, 2024 ZDHS (n = 12,133)

| Variable | Category | COR (95% CI) | AOR (95% CI) | p-value (AOR) |
|---|---|---|---|---|
| Age group | 15-24 years | 1.00 | 1.00 | — |
|  | 25-34 years | 0.79 (0.68-0.91) | 0.70 (0.59-0.82) | <0.001 |
|  | 35-49 years | 0.88 (0.76-1.01) | 0.65 (0.52-0.83) | <0.001 |
| Education | No education | 1.00 | 1.00 | — |
|  | Primary | 0.65 (0.54-0.78) | 0.76 (0.60-0.96) | 0.023 |
|  | Secondary | 0.73 (0.60-0.89) | 0.82 (0.63-1.06) | 0.120 |
|  | Higher | 0.43 (0.34-0.55) | 0.92 (0.63-1.35) | 0.686 |
| Marital status | Formerly married | 1.00 | 1.00 | — |
|  | Never married | 4.01 (3.17-5.07) | 2.20 (1.69-2.86) | <0.001 |
|  | In union | 4.39 (3.50-5.51) | 2.33 (1.90-2.86) | <0.001 |
| Wealth index | Poorest | 1.00 | 1.00 | — |
|  | Poorer | 0.86 (0.75-0.98) | 0.99 (0.84-1.16) | 0.865 |
|  | Middle | 0.77 (0.67-0.88) | 0.76 (0.63-0.91) | 0.004 |
|  | Richer | 0.73 (0.63-0.85) | 0.77 (0.61-0.97) | 0.031 |
|  | Richest | 0.66 (0.56-0.78) | 0.62 (0.45-0.86) | 0.004 |
| Parity | 0 children | 1.00 | 1.00 | — |
|  | 1-2 children | 1.80 (1.53-2.12) | 2.04 (1.64-2.54) | <0.001 |
|  | 3-4 children | 2.34 (1.99-2.75) | 2.25 (1.71-2.95) | <0.001 |
|  | 5+ children | 2.67 (2.27-3.14) | 3.05 (2.25-4.13) | <0.001 |
| Residence | Rural | 1.00 | 1.00 | — |
|  | Urban | 0.84 (0.74-0.96) | 0.80 (0.68-0.95) | 0.011 |
| Employment | Not working | 1.00 | 1.00 | — |
|  | Working | 0.73 (0.65-0.82) | 0.73 (0.64-0.84) | <0.001 |
| Fertility preference | Per unit increase | 1.19 (1.12-1.26) | 1.17 (1.10-1.24) | <0.001 |
| Distance to facility | No problem | 1.00 | 1.00 |  |
|  | Yes (problem) | 1.20 (1.09-1.33) | 0.89 (0.78-1.01) | 0.067 |
| Province | Central | 1.00 | 1.00 |  |
|  | Copperbelt | 1.09 (0.89-1.33) | 1.09 (0.84-1.40) | 0.524 |
|  | Eastern | 1.02 (0.83-1.25) | 1.01 (0.80-1.28) | 0.925 |
|  | Luapula | 1.06 (0.86-1.30) | 1.07 (0.83-1.37) | 0.618 |
|  | Lusaka | 1.42 (1.17-1.72) | 1.45 (1.13-1.86) | 0.003 |
|  | Muchinga | 1.21 (0.99-1.48) | 1.29 (1.00-1.68) | 0.053 |
|  | Northern | 1.32 (1.09-1.60) | 1.40 (1.11-1.77) | 0.005 |
|  | North-Western | 1.44 (1.16-1.78) | 1.46 (1.10-1.93) | 0.008 |
|  | Southern | 1.41 (1.17-1.70) | 1.51 (1.15-1.98) | 0.003 |
|  | Western | 1.13 (0.91-1.40) | 1.26 (0.97-1.63) | 0.086 |

COR = crude odds ratio; AOR = adjusted odds ratio; CI = confidence interval. Reference categories are shown with an odds ratio of 1.00. Estimates were derived from svyglm (quasibinomial family); province was included in the adjusted model. Statistical significance was set at p < 0.05.

Compared with women aged 15-24 years, the odds of unmet need were lower among women aged 25-34 years (AOR 0.70, 95% CI: 0.59-0.82) and 35-49 years (AOR 0.65, 95% CI: 0.52-0.83).

Women in the middle (AOR 0.76, 95% CI: 0.63-0.91), richer (AOR 0.77, 95% CI: 0.61-0.97) and richest (AOR 0.62, 95% CI: 0.45-0.86) wealth quintiles had lower odds than the poorest women, and women with primary education had lower odds than those with no education (AOR 0.76, 95% CI: 0.60-0.96). Working women (AOR 0.73, 95% CI: 0.64-0.84) and urban residents (AOR 0.80, 95% CI: 0.68-0.95) also had lower odds of unmet need. Each unit increase in fertility preference was associated with higher odds of unmet need (AOR 1.17, 95% CI: 1.10-1.24). Distance to a health facility was associated with unmet need in the crude analysis but not after adjustment (AOR 0.89, 95% CI: 0.78-1.01).

After adjustment for individual-level factors, women in Southern (AOR 1.51, 95% CI: 1.15-1.98), North-Western (AOR 1.46, 95% CI: 1.10-1.93), Lusaka (AOR 1.45, 95% CI: 1.13-1.86) and

Northern (AOR 1.40, 95% CI: 1.11-1.77) provinces had higher odds of unmet need than women in Central Province. Sensitivity analyses that retained partner’s fertility desire and decision-making autonomy with missing-category indicators did not materially change these estimates

## 4. Discussion

This analysis of the 2024 ZDHS found that more than one in four women of reproductive age in Zambia had an unmet need for family planning, that unmet need was geographically clustered with consistent hotspots in Lusaka, Southern and North-Western provinces, and that marital status, parity, age, household wealth, employment and place of residence were independently associated with unmet need. Because the analysis was cross-sectional, these relationships should be interpreted as associations rather than causal effects.

The burden observed is consistent with the continuing challenge of unmet need reported across sub-Saharan Africa (4, 13). Although our estimate is higher than the approximately 20% reported among married women in the 2018 ZDHS [2], the difference should be interpreted cautiously: our analysis included all women of reproductive age rather than married women only, and differences between survey rounds may also reflect measurement, fertility intentions and analytical procedures rather than a true change in unmet need. Previous Zambian studies have documented persistent inequalities in contraceptive use and unmet need across socioeconomic and demographic groups [3, 7]. Comparisons across ZDHS rounds should therefore use consistent definitions, denominators and analytical approaches.

The spatial findings add an important dimension by showing that unmet need is geographically patterned rather than randomly distributed. Similar spatial variation has been documented in Uganda and Ethiopia, where geographic concentration of unmet need has been linked to differences in population characteristics, socioeconomic conditions and access to reproductive health services [8, 9]. The consistency of the findings across Moran’s I, Getis-Ord Gi* and the scan statistic, together with the elevated adjusted odds in the same provinces, supports the conclusion that geographic context matters, although the three spatial methods are not independent because they rely on the same EA-level data. The location of the hotspots differs from earlier Zambian reports that emphasised Luapula and Eastern provinces [7], which may reflect change over time, different methods or different populations analysed. Spatial clustering does not establish that place itself causes unmet need, because clustered areas may share unmeasured social, economic or health-service characteristics.

The concentration of unmet need in Lusaka is notable given that urban residence was associated with lower odds at the individual level. These findings are not necessarily contradictory, because an urban classification can conceal substantial heterogeneity. Women living in the same city may experience very different access to family planning depending on poverty, settlement characteristics, mobility, information, and service availability [14, 15], and proximity to an urban centre does not necessarily translate into adequate family planning information or effective access in informal settlements [16]. The Lusaka cluster may therefore point to intra-urban, possibly peri-urban, inequalities. Because settlement type and facility accessibility were not measured, this explanation remains a hypothesis for further study.

Younger women had higher odds of unmet need than older women, consistent with evidence from sub-Saharan Africa that young women face distinctive barriers to contraceptive access and use [5]. Qualitative studies have identified concerns about confidentiality, stigma, provider attitudes, limited information, and fear of judgement as barriers among adolescents and young women [17, 18]. The higher odds among women in union and never-married women, relative to formerly married women, are consistent with variation by relationship status reported in Zambia and elsewhere [7, 13]. Partner communication, partner opposition, and women’s decision-making autonomy may contribute [2], but these factors could not be examined here because the relevant variables had high levels of missing data.

The dose-response relationship between parity and unmet need is consistent with evidence that reproductive history shapes family planning requirements, particularly as women seek to space or limit further births [4, 19]. Women with several children may also have experienced contraceptive discontinuation, side effects, or dissatisfaction with methods. Contacts with maternal, postpartum and child health services may therefore offer important opportunities to identify and address unmet need. The lower odds among wealthier, educated and employed women are consistent with the role of enabling resources in access to health services [11] and with documented socioeconomic inequalities in contraceptive use in Zambia [3]. The loss of association with distance to a health facility after adjustment suggests that its crude effect was explained by correlated factors such as residence and wealth.

### 4.1 Study strengths and limitations

The main strengths of this study are the use of recent, nationally representative, geo-referenced data; analyses that accounted for the complex survey design; and the combination of three complementary spatial methods with survey-weighted regression. Several limitations should be considered. First, the cross-sectional design precludes assessment of temporality and causality. Second, unmet need is derived from self-reported fertility intentions and contraceptive behaviour and may be affected by recall and social desirability bias. Third, we excluded partner fertility desire and decision-making autonomy because of high missingness, so residual confounding by relational factors is possible, although sensitivity analyses did not materially change the estimates. Fourth, parity, marital status, and fertility preference are closely related to the definition of unmet need, so their associations should be interpreted with caution. Fifth, the random displacement of DHS cluster coordinates may have misplaced some EAs and attenuated local clustering; EA-level prevalence is based on a small number of women per EA, and the scan statistic did not incorporate survey weights or adjust for covariates; area-level patterns should not be interpreted at the individual level. Finally, a single survey round does not allow trends in the spatial pattern to be assessed.

## 4.2 Conclusion

Unmet need for family planning remains a substantial public health concern in Zambia. It is geographically clustered, with hotspots in Lusaka, Southern and North-Western provinces, and is concentrated among younger, higher-parity and socioeconomically disadvantaged women. National averages alone are therefore insufficient to guide family planning programmes.

We recommend that the Ministry of Health and its partners: (i) prioritise the identified hotspot areas, including peri-urban Lusaka, Southern and North-Western provinces, when allocating family planning resources and outreach; (ii) strengthen confidential, youth-friendly contraceptive services for adolescents and young women; (iii) integrate family planning counselling into maternal, postpartum and child health contacts to reach higher-parity women; and (iv) reduce financial barriers to contraception for the poorest households. Future research should link survey data with information on health-facility readiness, contraceptive availability, service quality, partner dynamics and community characteristics to explain the observed spatial inequalities.

## Data Availability

No data was generated by this study. The study used publicly available data from the 2024 Zambia Demographic and Health Survey (ZDHS), implemented by the Zambia Statistics Agency in partnership with the Ministry of Health, with technical assistance from ICF through The DHS Program. The datasets are available from the DHS Program at https://dhsprogram.com/data/available-datasets.cfm after free registration and approval of a dataset access request. The authors had no special access privileges to these data.

## Data availability statement

Third-party data were obtained for this study from the DHS Program. Data may be requested from the DHS Program after creating an account and submitting a concept note (https://dhsprogram.com/data/Access-Instructions.cfm). Data for this study were analysed from the 2024 Zambia Demographic and Health Survey (ZDHS). The authors confirm that interested researchers would be able to access these data in the same manner as the authors, and that the authors had no special access privileges that others would not have.

## Acknowledgments

We thank the University of Zambia School of Public Health for an enabling academic environment, the Zambia Statistics Agency and ICF for open access to the 2024 ZDHS data, and the Zambia National Public Health Institute for logistical support.

## Author contributions

Conceptualization: Philemon Sibeso

Data curation:Philemon Sibeso

Formal analysis: Philemon Sibeso.

Methodology: Philemon Sibeso

Software: Philemon Sibeso

Supervision: Nasson Tembo

Visualization: Philemon Sibeso

Writing original draft: Philemon Sibeso

Writing review & editing: Proffessor cholwe jacobs

